# Improving prediction of survival and progression in metastatic non-small cell lung cancer following immunotherapy through machine learning of circulating tumor DNA dynamics

**DOI:** 10.1101/2023.10.24.23297462

**Authors:** Haolun Ding, Min Yuan, Yaning Yang, Xu Steven Xu

**Author notes:** Corresponding author: Min Yuan and Xu Steven Xu. HD and MY contributed equally.

## Abstract

**Objectives:** To use modern machine-learning approaches to enhance and automate the feature extraction from the longitudinal ctDNA data and to improve the prediction of survival and disease progression, risk stratification, and treatment strategies for patients with 1L NSCLC.

**Methods:** Using IMpower150 trial data on untreated metastatic non-small cell lung cancer patients treated with atezolizumab and chemotherapies, we developed a machine-learning algorithm to extract predictive features from ctDNA kinetics, improving survival and progression prediction. We analyzed kinetic data from 17 ctDNA summary markers, including cell-free DNA concentration, allele frequency, tumor molecules in plasma, and mutation counts. Our machine-learning workflow (FPCRF) involved functional principal component analysis (FPCA) for automated feature extraction, random forest and bagging ensemble algorithms for feature selection, standard PCA for dimension reduction, and Cox proportional-hazards regression for survival analysis. The dataset was divided into training and test cohorts in the same way as a previous study.

**Results:** 398 patients with ctDNA data (206 in training, 192 in validation) were analyzed. Our machine-learning models automated feature extraction, excelling in predicting overall survival (OS) and progression-free survival (PFS) at different landmarks. In identical train-test cohorts, our models outperformed existing ones using handcrafted ctDNA features, raising OS c-index to 0.72 and 0.71 from 0.67 and 0.63 for C3D1 and C4D1, and substantially improving PFS to ∼0.65 from the previous 0.54 - 0.58, a 12-20% increase. Our model enhanced risk stratification for NSCLC patients, achieving clear OS and PFS separation (e.g., on C3D1, HR: 2.65 [95%CI: 1.78–3.95, P < 0.001] for high vs. intermediate risk, 2.06 [95%CI: 1.29–3.29, P = 0.002] for intermediate vs. low risk; and PFS HR: 2.04 [95%CI: 1.41–2.94, P < 0.001], 1.56 [95%CI: 1.07–2.27, P = 0.02]). Distinct patterns of ctDNA kinetic characteristics (e.g., baseline ctDNA markers, depth of ctDNA responses, and timing of ctDNA clearance, etc.) were revealed across the risk groups. Rapid and complete ctDNA clearance appears essential for long-term clinical benefit.

**Conclusions:** Our machine-learning approach offers a novel tool for analyzing ctDNA kinetics, extracting critical features from longitudinal data, improving our understanding of the link between ctDNA kinetics and progression/mortality risks, and optimizing personalized immunotherapies for 1L NSCLC.

**Research in context:** *Evidence before this study:* The longitudinal dynamics of ctDNA are showing promise as a biomarker for treatment outcomes and monitoring. However, despite of recent advances of machine learning, very limited applications have been reported in using machine learning-based approaches to analyze the longitudinal ctDNA data, improve the prediction of clinical outcomes, and refine the risk stratifications. We searched PubMed on Oct 8, 2023 for peer-reviewed, English-language journal and conference articles using the terms (“ctDNA”) AND (“deep learning” OR “machine learning” OR “artificial intelligence”). Fifty-nine (59) search results were found. After systematical review of these search results, we found only 4 research studies where longitudinal ctDNA dynamic/kinetic data were analyzed using machine-learning models to predict patient outcomes. These studies focused on building models using handcrafted features of ctDNA dynamics such as on-treatment ctDNA levels and early ctDNA changes and clearance, etc. So far, no studies have utilized machine- or deep-learning models to extract features from longitudinal ctDNA dynamics to inform and predict cancer patient outcomes.

*Added value of this study:* We developed a machine-learning algorithm to predict survival and disease progression using ctDNA data from the Impower150 trial on untreated metastatic non-small cell lung cancer patients receiving atezolizumab and chemotherapy. Our machine-learning models automatically extract informative features from longitudinal ctDNA dynamics, outperforming existing models based on handcrafted features in predicting overall survival and progression-free survival at various time points. They improved risk stratification and identified crucial ctDNA kinetic characteristics in 1L NSCLC, revealing the importance of rapid and complete ctDNA clearance for long-term clinical benefit.

*Implications of all the available evidence:* Machine-learning models can automatically extract prognostic features from longitudinal ctDNA dynamic trajectories, enable refined risk stratification and prediction of clinical outcomes, and thereby enhance ctDNA data’s utility in clinical patient care and personalized treatment.

## Introduction

Circulating tumor DNA (ctDNA) holds immense importance and utility in the field of oncology and personalized medicine [1]. This emerging, non-invasive biomarker consists of fragments of DNA shed into the bloodstream by cancer cells, allowing for the detection and monitoring of cancer without the need for invasive procedures like tissue biopsies [2]. Moreover, longitudinal ctDNA measurements enable the real-time monitoring of treatment response, the early detection of minimal residual disease, and the identification of emerging resistance mechanisms [3, 4]. Previous analyses have shown the significant contributions of longitudinal information of ctDNA to risk estimation [4–7], offering a promising avenue for improved outcomes for cancer patients.

Currently, risk stratification models for longitudinal biomarkers are often based on handcrafted features from the longitudinal trajectories [8, 9]. Most recently, a machine learning model for ctDNA was developed using multiple handcrafted ctDNA metrics, such as ctDNA levels and number of alterations at certain times, change in mutations from baseline, etc. to predict clinical outcomes [10]. This model suggests that measuring ctDNA dynamics during treatment can improve patient risk stratification and may allow early differentiation between competing therapies during clinical trials.

Due to the complexity of on-treatment biomarker trajectories, more advanced modeling approaches may be required to capture complex relationships among different biomarkers over time and between biomarker changes and clinical outcomes accurately. In recent years, there has been a surge in the adoption of machine and deep learning methods, many of which leverage longitudinal or time-series data for outcome prediction [11–15]. Machine learning models can capture temporal dependencies, detect trends, and reveal hidden patterns and insights within complex longitudinal data. It has been shown that extracting features from longitudinal data using machine learning may be a powerful approach for reducing the dimensionality of the data and identifying meaningful features and relationships within the longitudinal data, allowing us to make better predictions, classify outcomes, or understand the underlying dynamics of disease progression [16].

Based on the data from the IMpower150 trial [17], this study aimed to overcome current limitations in longitudinal ctDNA modeling based on handcrafted features and utilize machine-learning algorithms for enhancing feature extraction from longitudinal ctDNA kinetics automatically to improve clinical outcome predictions and optimize risk stratifications of patients with untreated metastatic non-squamous non-small-cell lung cancer (NSCLC) following immuno- and chemotherapy.

## Materials and Methods

### Patient Data Set

The analysis in this study utilized data from the IMpower150 trial [17], which was a phase 3 clinical trial that evaluated the efficacy and safety of adding atezolizumab, an immune checkpoint inhibitor, to a combination of bevacizumab, carboplatin, and paclitaxel in patients with metastatic non-squamous NSCLC who had not received prior chemotherapy. The trial had three treatment arms: ACP (atezolizumab plus carboplatin plus paclitaxel), BCP (bevacizumab plus carboplatin plus paclitaxel), and ABCP (atezolizumab plus BCP). The trial included patients with and without EGFR or ALK genetic mutations, as well as patients with different levels of PD-L1 expression and tumor-infiltrating effector T cells. These patients were divided into three groups in a 1:1:1 ratio, and treated with either ACP, or BCP, or ABCP in eight 3-week cycles. The trial demonstrated that adding atezolizumab to bevacizumab plus chemotherapy improved patient outcomes across different biomarker subgroups and provided a new treatment option for this patient population. The trial protocol received ethical approval from site-specific ethics committees, adhering to the Declaration of Helsinki and Good Clinical Practice guidelines. Prior to enrollment, patients provided voluntary written consent without compensation. The trial is registered on www.ClinicalTrials.gov under identifier NCT02366143.

ctDNA was collected from 566 patients using a 330 kb custom assay targeting 311 genes [10]. Longitudinal ctDNA measurements were taken at five timepoints: baseline (BL), cycle 2 day 1 (C2D1; Week 3), cycle 3 day 1 (C3D1; Week 6), cycle 4 day 1 (C4D1; Week 9), and cycle 8 day 1 (C8D1; Week 21). Multiple ctDNA summary metrics, including mean, median, and maximum values, were used to characterize ctDNA levels, such as cell-free DNA concentration (cfDNA), allele frequency (AF), tumor molecules per ml plasma (TMPMP), and mutation counts. The analysis also included ctDNA levels for a subset of known or likely pathogenic mutations. In total, 17 ctDNA summary markers were derived and used (Table S1). After excluding patients with missing data, our analysis included 398 patients. The clinical outcomes of interest in this analysis were overall survival (OS) which refers to the duration from randomization to death for any reason and progression-free survival (PFS), assessed by investigators according to RECIST v1.1 criterion.

### Machine learning of ctDNA trajectories and survival

Figure 1 outlines the workflow for our analysis. This process involved reducing the dimensionality of longitudinal ctDNA data and feature extraction, feature selection, survival analysis for clinical outcome prediction, risk classification, and identification of critical ctDNA kinetic characteristics informing patient’s risk.

**Figure 1.**
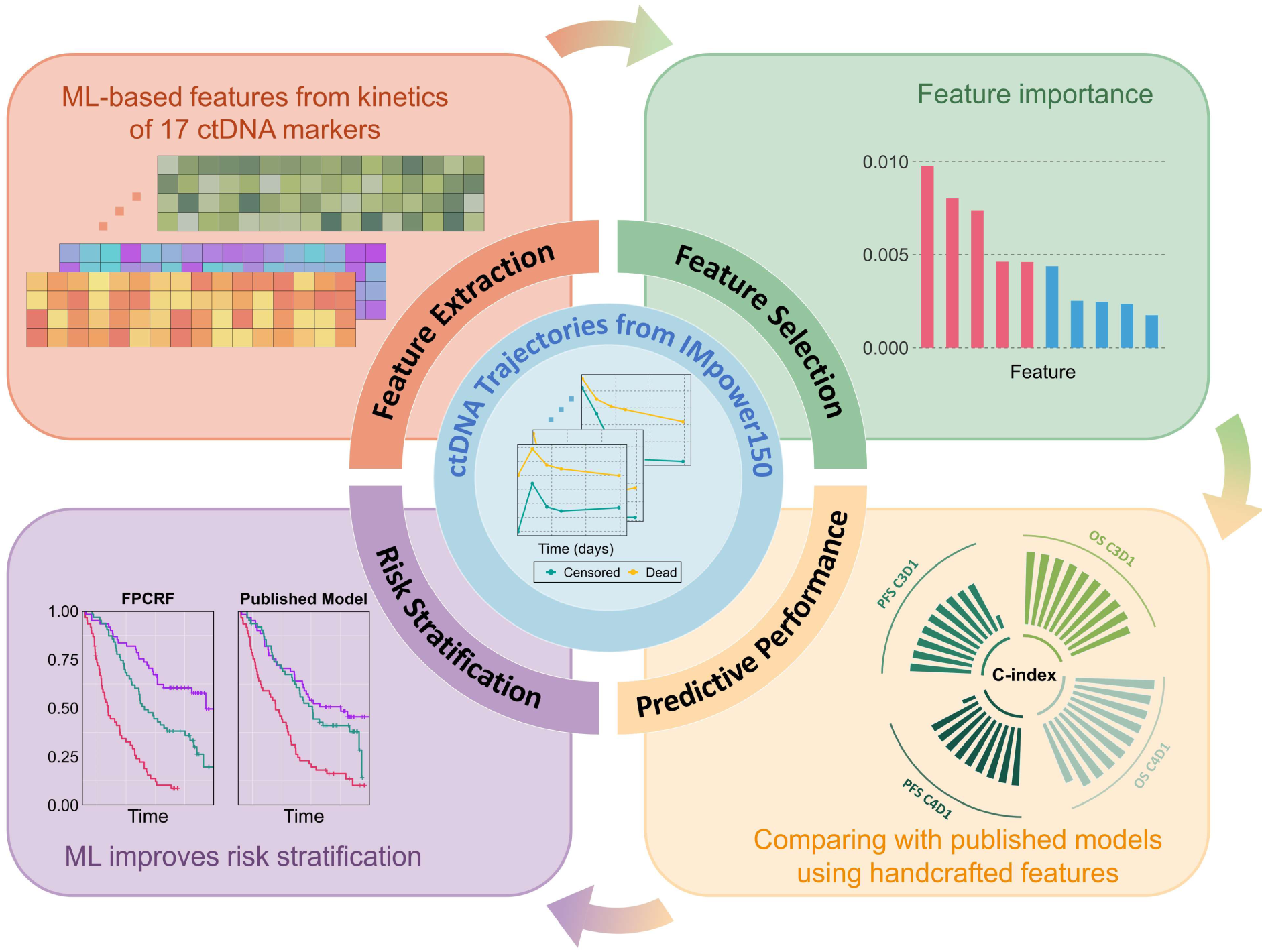
Workflow of Machine Learning of Longitudinal Circulating Tumor DNA and Clinical Outcomes

#### Univariate workflow

Functional Principal Component Analysis (FPCA) was used to extract features from each summary ctDNA marker trajectory up to different landmark time points (C3D1 and C4D1). It employs a low-dimensional representation of data through Karhunen-Loève decomposition, transforming each trajectory into a linear combination of functional principal components (FPCs) [18]. FPCs are orthogonal eigenfunctions capturing trend variations via Mercer’s Theorem [19]. Using FPCA, we obtained low-rank representations of each ctDNA marker’s trajectory for each patient. The first two FPCs were selected for feature extraction (Figure S3) and survival analysis using Cox proportional-hazards regression [20]. Model fitting was performed with the “coxph” function in the R survival package [21].

#### Multivariate workflow (FPCRF)

Trajectory features extracted using FPCA from the 17 ctDNA summary markers were combined. Since correlations were observed among the 17 ctDNA markers (Figure 2B and 2C) and the features (Figure S4), these features were screened using random forest (RF) and bagging ensemble algorithms with conditional inference trees as basic learners [22–24]. The top important features were selected using “cforest” and “varimp” functions in R “party” package [22] (Figure S5) and then PCA was applied to further reduce dimensionality and collinearity, retaining the final feature matrix capturing 99% of variability of the ctDNA data. This final feature matrix was used as the input in the Cox regression models. This workflow is referred to as FPCRF thereafter in the manuscript.

**Figure 2.**
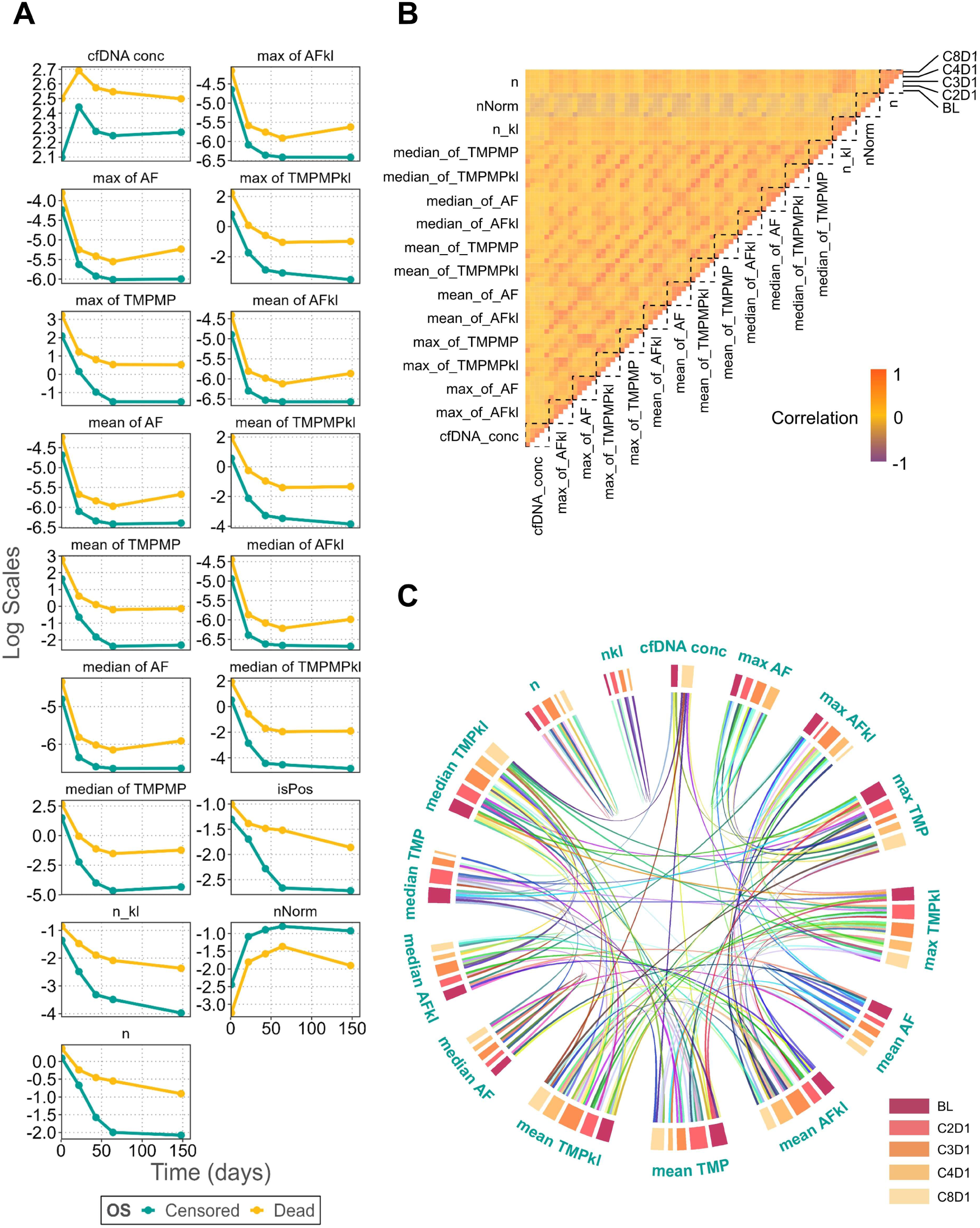
Evolution and correlation of longitudinal ctDNA markers over time. A. mean profiles for censored and decreased patients; B. Correlation of longitudinal ctDNA markers in each period.; C. Network analysis to depict relationships among ctDNA dynamic markers.

### Model training and validation

To ensure a fair comparison with published models, the dataset was split into training and test cohorts in the same way as a previous study [10]. Concordance index (c-index) [25] was used to evaluate the predictive performance between FPCRF and the published method.

### Model-based risk stratification

Risk stratification was implemented by dividing patients into three subgroups according to tertiles of the predictive risk. We compared OS and PFS probabilities using Kaplan-Meier plots for subgroups identified at C3D1 and C4D1 landmark time points. The log rank test was used to evaluate the statistical significance among subgroups and c-index was used to evaluate performance of the risk stratification.

## Results

The training dataset included 206 patients while 192 patients were included in the test data. Patient demographics are summarized elsewhere [10, 17]. The minimum duration of follow-up for OS was approximately 1.87 and 1.94 months, in the training and test data, respectively (Figure S1). A total of 144 (69.9%) and 132 (68.8%) of the patients in the training and test data had died. The overall survival was similar in the training data compared to the test data (median, 20.52 months vs. 21.83 months). The minimum duration of follow-up was 1.18 months for PFS. 183 (88.8%) and 175 (91.2%) had disease progression or died in the training and test data, respectively. Progression-free survival was virtually identical in the training and test data (median, 7.84 months vs. 8.07 months, respectively).

### Dynamics of different ctDNA Summary Markers

In Figure 2A and Figure S2, we display the average trajectories of 17 ctDNA markers (Table S1) for patients who either survived or deceased. These average trajectories reveal distinct patterns between the two groups. Most notably, the deceased group generally displayed higher levels of ctDNA markers compared to the censored/survival group. In Figure 2B, we observe strong correlations in the kinetics of the 17 distinct ctDNA markers. Employing a correlation threshold of 0.8, we constructed a network diagram that illustrates the relationships among these ctDNA markers at different time points, providing further insights into their kinetic relationships.

### Predictive performance of univariate FPCA and multivariate FPCRF

On the independent test data, our model (FPCRF) demonstrated outstanding predictive performance for both OS and PFS on C3D1 and C4D1 (Figure 3 and Table S2). When we trained our model on the same dataset (206 patients) as a previously published model and tested it on the identical test dataset (192 patients), FPCRF substantially outperformed the published model using handcrafted ctDNA features. Specifically, it improved the c-index for OS by 6.4% on C3D1 (c-index = 0.72 for FPCRF vs. 0.67 for the published model) and 13.2% on C4D1 (c-index = 0.71 for FPCRF vs. 0.63 for the published model). Even more remarkable was the improvement in predicting PFS. FPCRF achieved a c-index of 0.65 on both C3D1 and C4D1, respectively, which were 20.3% and 12.1% higher than those of the published model (c-indices of 0.54 and 0.58 on C3D1 and C4D1, respectively).

**Figure 3.**
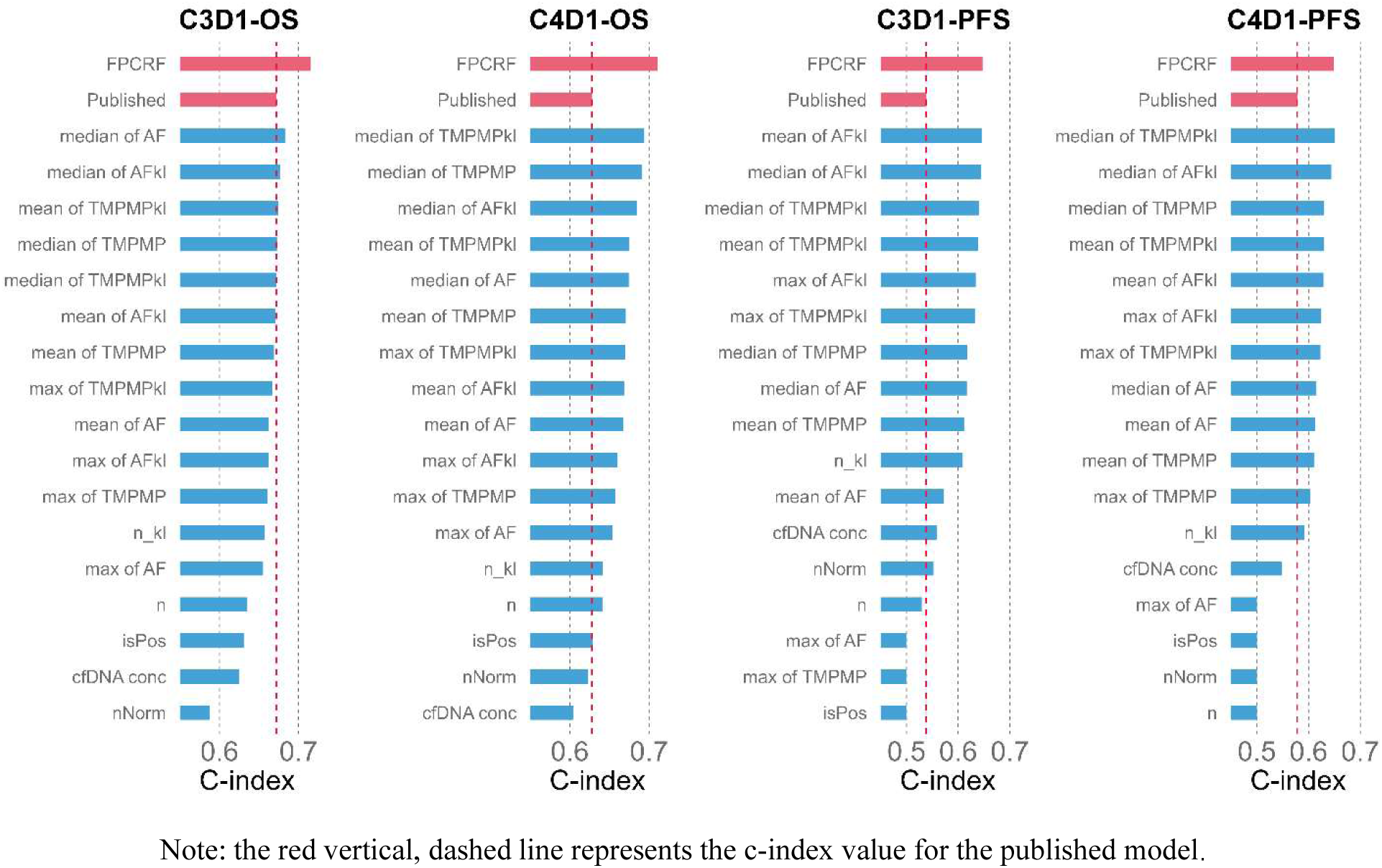
Comparison of Predictive performance of univariate FPCA, FPCRF, and published models.

Notably, univariate FPC features extracted from the ctDNA kinetics of a single marker, also exhibit superior performance compared to the previously published model that was developed from handcrafted ctDNA features from longitudinal data of all 17 ctDNA markers (Figure 2 and Table S2). Specifically, on C3D1, both median AF and AFkl kinetics produced slightly higher c-indices (0.68) for predicting OS than the published model [10]. Furthermore, machine-learning derived features for several other ctDNA markers, including mean, median, and maximum values of TMPMPkl, as well as the mean and median values of TMPMP and the mean of AFkl, offered equivalent predictive performance to the existing model.

On C4D1, univariate machine-learning features extracted from most ctDNA markers (14 out of 17) substantially outperformed the existing model. The only exceptions were the predictions based on the kinetic features from ctDNA positivity status, cfDNA concentration, and nNorm, which were slightly suboptimal compared to the existing model. Similarly, in predicting PFS, the vast majority of ctDNA markers in univariate analysis demonstrated notably improved predictive performance in comparison to the existing model. These results underscore the prognostic value of machine-learning extracted univariate FPC features and highlight their superiority over the handcrafted features employed in the published model.

### Risk stratification and clinical interpretation

We risk stratified the NSCLC patients in the test cohort into three risk groups (low, intermediate, and high risk) based on the tertiles of the risk determined by the FPCRF models. The FPCRF ctDNA classifiers consistently demonstrated improved and more statistically significant distinctions for OS (Figure 4) compared to the published model.

**Figure 4.**
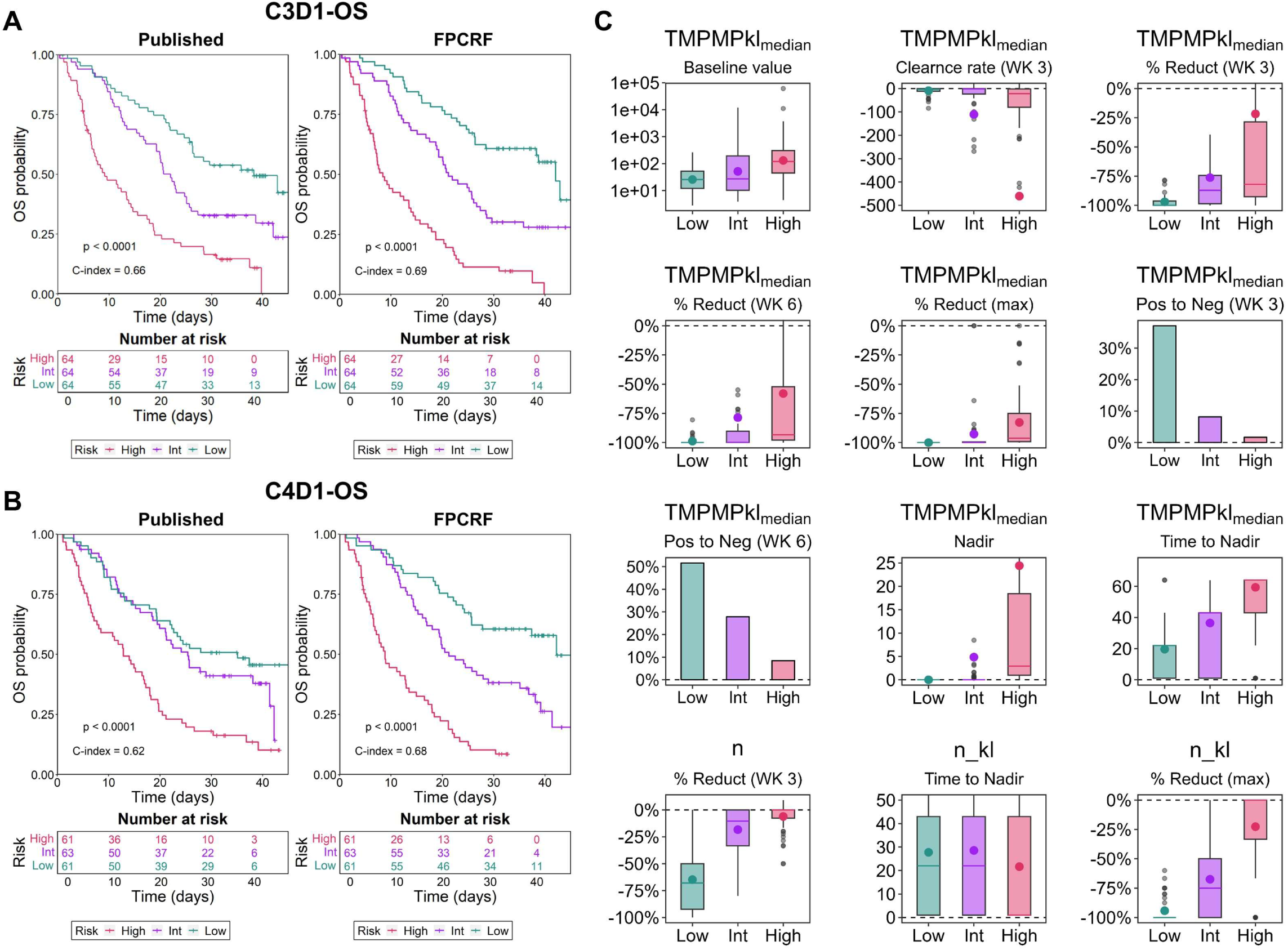
Risk stratification for overall survival based on machine learning models and published models (A: C3D1 OS and B: C4D1 OS); C: selected ctDNA kinetic characteristics of different risk groups

At C3D1, although both the published model and the FPCRF model effectively separated the OS curves of the three risk groups (Figure 4A), the FPCRF stratification emerged as a stronger predictor of OS. Specifically, for intermediate risk compared to low risk, the Hazard Ratio (HR) was 2.06 [95%CI: 1.29–3.29, *P* = 0.002] for FPCRF and 1.75 [95%CI: 1.11–2.76, *P* = 0.016] for the published model. For high risk versus intermediate risk, the HR was 2.65 [95%CI: 1.78–3.95, *P* < 0.001] for FPCRF and 2.19 [95%CI: 1.47–3.26, *P* < 0.001] for the published model. The c-index for FPCRF stratification was 0.69, whereas it was 0.66 for the published model.

On C4D1, the superiority of FPCRF stratification for OS over the published model became even more evident (Figure 4B). For intermediate risk compared to low risk, the estimated HR was 2.07 [95%CI: 1.27–3.36, *P* = 0.004] for FPCRF and 1.27 [95%CI: 0.80–2.03, *P* = 0.3] for the published model. Meanwhile, for high risk versus low risk, the estimated HR was 5.72 [95%CI: 3.51–9.31, *P* < 0.001] for FPCRF and 2.80 [95%CI: 1.80–4.36, *P* < 0.001] for the published model. Consequently, the FPCRF stratification significantly improved the separation between low- and intermediate-risk groups compared to the published model. The c-index for FPCRF stratification was 0.68, compared to 0.62 based on the published model.

The FPCRF classifiers also displayed a notable advantage in separating and classifying PFS compared to the published model (Figure 5). On C3D1, the published model struggled to differentiate the PFS curves between the high- and intermediate-risk groups (HR = 1.07 [95%CI: 0.74–1.53, P = 0.7]), although some distinction was seen between high- and low-risk groups (HR = 3.01 [95%CI: 2.04–4.44, P < 0.001]) (Figure 5A). Conversely, the FPCRF classifier demonstrated clear separation of PFS curves for all three risk groups. The estimated HR for the high- and intermediate-risk groups was 2.04 [95%CI: 1.41–2.94, P < 0.001], and for the intermediate- vs. low-risk groups, it was 1.56 [95%CI: 1.07–2.27, P = 0.02]. The overall P value for the FPCRF classifier was < 0.0001, with a c-index of 0.62, compared to 0.014 and 0.56 for the published model.

**Figure 5.**
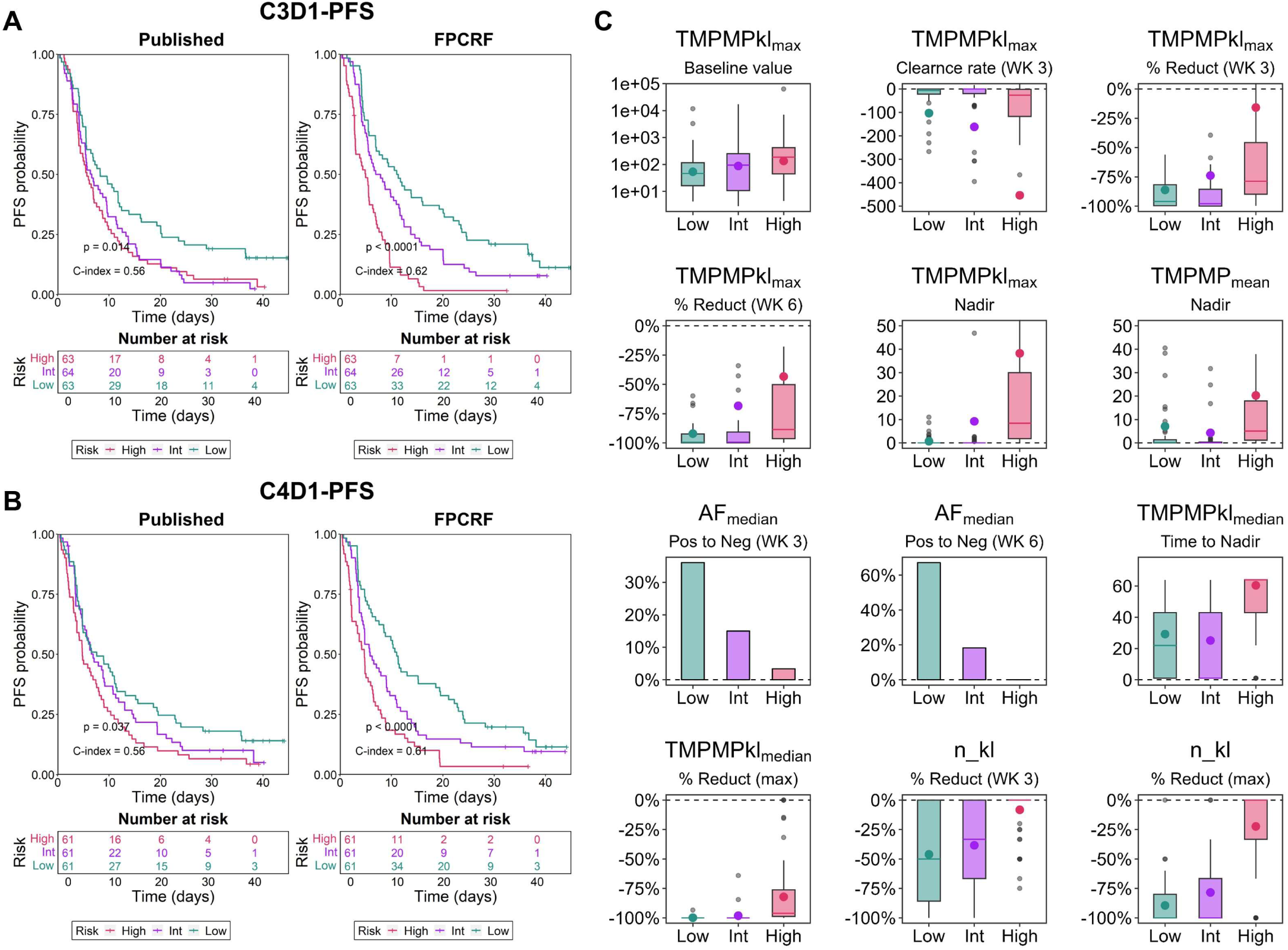
Risk stratification for PFS based on machine learning models and published models (A: C3D1 PFS and B: C4D1 PFS); C: selected ctDNA kinetic characteristics of different risk groups

Similar patterns were observed at C4D1. The published PFS model’s risk stratification failed to provide clear separations among the three risk groups (for high- vs. intermediate-risk groups, HR = 1.31 [95%CI: 0.91–1.90, P = 0.15]; and for intermediate- vs. low-risk groups, HR = 1.24 [95%CI: 0.85–1.81, P = 0.3]). In contrast, the FPCRF classifiers effectively distinguished PFS curves (for high- vs. intermediate-risk groups, HR = 1.62 [95%CI: 1.12–2.35, P = 0.011]; and for intermediate- vs. low-risk groups, HR = 1.48 [95%CI: 1.01–2.17, P = 0.043]) (Figure 5B). The overall P value for the FPCRF classifier was < 0.0001, and its c-index was 0.61, compared to 0.037 and 0.56 for the published model.

To gain deeper insights into the ctDNA kinetic attributes within each risk group, we conducted a comparative analysis of key kinetic metrics across these groups (Figure 4C and Figure 5C). Notably, Figure 4C and Table S3 shows that baseline ctDNA markers, including metrics like the baseline median, mean, and maximum profiles of TMPMP and TMPMPkl, along with cfDNA concentrations, generally showed an increase as the risk of mortality escalated across the risk categories. Furthermore, the baseline prevalence of ctDNA positivity tended to rise in patients categorized as low, intermediate, and high risk.

Interestingly, the high-risk group seemingly exhibited a more rapid rate of ctDNA clearance (i.e., the slope of ctDNA decline) at both Week 3 (C2D1) and Week 6 (C3D1) compared to the low- risk group. This phenomenon could be attributed to the infrequent collection of ctDNA samples (every 3 weeks), which limited the accuracy of rate calculations for ctDNA decline. For instance, patients in the low-risk group might have achieved ctDNA clearance much earlier than Week 3.

Moreover, as indicated in Figure 4C, at Week 3, nearly all low-risk patients achieved complete clearance of ctDNA (a 96% reduction) from their circulation, whereas high-risk patients exhibited only an average of ∼17% reduction, with few high-risk patients reaching complete clearance by Week 3, suggesting a slow clearance of ctDNA in high-risk patients. In line with these findings, patients with a higher risk profile tended to have a higher nadir and required a longer time to reach that nadir, while all low-risk patients cleared ctDNA completely, for instance, in terms of median TMPMPkl, with an average time to nadir of 3 weeks (21 days). These results emphasize the importance of swiftly and thoroughly removing ctDNA from circulation. Similarly, greater reductions in mutations (n and n_kl) were observed in patients with lower risk (i.e., low- vs intermediate- vs high-risk patients). However, no clear trend was observed for the time it took to reach the nadir of mutations across different risk categories.

Similar patterns of ctDNA kinetic characteristics were observed for PFS-based stratifications (Figure 5C and Table S4). High-risk patients with a higher likelihood of progression are generally distinguishable from those with median and low risk based on various ctDNA kinetic metrics. However, the predictive value of cfDNA, nNorm, and n (number of mutations) for progression is limited, although n_kl (mutations in known or likely pathogenic genes) exhibits better predictive capabilities. Time to nadir appears to be less predictive of PFS compared to OS.

## Discussion

Longitudinal ctDNA data involve complex temporal correlations, between-patient variation, as well as associations among different ctDNA summary markers evolution over time. Modeling on- treatment ctDNA trajectories, particularly various ctDNA marker dynamics simultaneously can be challenging, given different distributions and evolutions of different markers. Currently, machine-learning models for ctDNA longitudinal data have been mainly using handcrafted features for ctDNA dynamics such as on-treatment ctDNA levels and early ctDNA changes and clearance, etc [10]. Our study aimed to use modern machine-learning approaches to enhance and automate the feature extraction from the longitudinal ctDNA data and to improve the prediction of survival and disease progression as well as risk stratification for patients with 1L NSCLC.

Using the same training and testing datasets from the Impower 150 trial, our FPCRF models substantially surpassed the existing models that were constructed with handcrafted features of ctDNA trajectories, achieving an average of 9.8% and 16.2% higher c-index in prediction OS and PFS, respectively. In addition, univariate analysis revealed the superiority of machine-learning based features from single ctDNA marker trajectories over published models using handcrafted features of multiple ctDNA markers. The superior predictive performance suggested that machine-learning based models could effectively extract critical prognostic information from the complex ctDNA dynamic data, capturing intricate relationships and time-varying patterns inherent in the ctDNA longitudinal data and providing excellent predictions for both OS and PFS of 1L untreated metastatic NLCSC following ICI and chemotherapy combinations. To our knowledge, this is the first study attempting to automate the feature extraction from longitudinal ctDNA dynamic data using machine-learning algorithms to improve the prognostication of survival outcomes and risk stratification of 1L metastatic NLCSC patients.

We stratified the mNSCLC patients into 3 risk groups (low, intermediate, and high risk) according to the model-predicted risk. The NSCLC patients in different risk groups demonstrated distinct survival outcomes as well as ctDNA kinetic characteristics. Our analysis indicates that clearance ctDNA from the circulation rapidly and thoroughly is critical. All the identified low risk patients had completely eliminated ctDNA from their system within 3 weeks. In contrast, intermediate- and high-risk patients were much less likely to achieve the ctDNA clearance status during the first 3 weeks. Additionally, it took much longer time for intermediate- and high-risk patients to achieve their nadir compared to the low-risk patients. The importance of the early ctDNA clearance may shed lights on future treatment strategies and dose selection in the 1L metastatic NSCLC.

Current ctDNA dynamics research also has limitations. Sparse sample collection may hamper understanding of ctDNA’s nonlinear nature over time, hindering accurate clearance rate evaluation and precise kinetic characterization [26, 27]. Future studies could optimize ctDNA sampling design for enhanced insights, better capturing early changes and progression [28]. Moreover, application of sophisticated models may help to improve the kinetic analysis for longitudinal ctDNA data that often include irregular and missing observations [29]. Overcoming these limitations will advance our comprehension of ctDNA dynamics and its clinical applications. Additionally, deep learning, specifically combining RNNs with CNNs or attention mechanisms, can enhance feature extraction [30–32], particularly for complex, high-dimensional longitudinal data. Future research may be warranted.

Taken together, the machine-learning-based approach is particularly valuable in studies of kinetics of ctDNA, allowing researchers to extract essential features from longitudinal ctDNA data, enhancing our ability to understand and model ctDNA dynamic processes, make informed predictions of patient outcomes, risk-stratify cancer patients, and optimize the patient care and treatment strategies for 1L NSCLC.

## Data Availability

All clinical and ctDNA data for IMpower150 are deposited to the European Genome-Phenome Archive under accession number EGAS00001006703. Qualified researchers may request access to individual patient-level data through the clinical study data request platform (https://vivli.org/).

## ACKNOWLEDGMENT

This work was partially supported by the Natural Science Foundation of Anhui Province (No. 2008085MA09 and 2022AH050703), the National Natural Science Foundation of China (No. 11671375).

## References

1. Sanz-Garcia, E., et al., Monitoring and adapting cancer treatment using circulating tumor DNA kinetics: Current research, opportunities, and challenges. SCIENCE ADVANCES, 2022. 8(4).

2. Nabet, B.Y., et al., Noninvasive Early Identification of Therapeutic Benefit from Immune Checkpoint Inhibition. CELL, 2020. 183(2): p. 363–376.

3. Anagnostou, V., et al., Dynamics of Tumor and Immune Responses during Immune Checkpoint Blockade in Non-Small Cell Lung Cancer. CANCER RESEARCH, 2019. 79(6): p. 1214–1225.

4. Lv, J.W., et al., Liquid biopsy tracking during sequential chemo-radiotherapy identifies distinct prognostic phenotypes in nasopharyngeal carcinoma. NATURE COMMUNICATIONS, 2019. 10: p.3941.

5. Powles, T., et al., ctDNA guiding adjuvant immunotherapy in urothelial carcinoma. NATURE, 2021. 595(7867): p. 432–437.

6. Moding, E.J., et al., Circulating tumor DNA dynamics predict benefit from consolidation immunotherapy in locally advanced non-small-cell lung cancer. NATURE CANCER, 2020. 1(2): p. 176–183.

7. Bratman, S.V., et al., Personalized circulating tumor DNA analysis as a predictive biomarker in solid tumor patients treated with pembrolizumab. NATURE CANCER, 2020. 1(9): p. 873–881.

8. Xu, X.S., et al., Correlation between Prostate-Specific Antigen Kinetics and Overall Survival in Abiraterone Acetate-Treated Castration-Resistant Prostate Cancer Patients. CLINICAL CANCER RESEARCH, 2015. 21(14): p. 3170–3177.

9. Yan, X.Y., et al., Early M-Protein Dynamics Predicts Progression-Free Survival in Patients With Relapsed/Refractory Multiple Myeloma. CTS-CLINICAL AND TRANSLATIONAL SCIENCE, 2020. 13(6): p. 1345–1354.

10. Assaf, Z.J.F., et al., A longitudinal circulating tumor DNA-based model associated with survival in metastatic non-small-cell lung cancer. NATURE MEDICINE, 2023. 29(4): p. 859–868

11. Netterberg, I., et al., A PK/PD Analysis of Circulating Biomarkers and Their Relationship to Tumor Response in Atezolizumab-Treated non-small Cell Lung Cancer Patients. CLINICAL PHARMACOLOGY & THERAPEUTICS, 2019. 105(2): p. 486–495.

12. Jin, C., et al., Predicting treatment response from longitudinal images using multi-task deep learning. NATURE COMMUNICATIONS, 2021. 12(1): p. 1851.

13. Li, F.L., et al., Deep learning-based predictive biomarker of pathological complete response to neoadjuvant chemotherapy from histological images in breast cancer. JOURNAL OF TRANSLATIONAL MEDICINE, 2021. 19(1): p. 348.

14. Gavrilov, S., et al., Longitudinal Tumor Size and Neutrophil-to-Lymphocyte Ratio Are Prognostic Biomarkers for Overall Survival in Patients With Advanced Non-Small Cell Lung Cancer Treated With Durvalumab. CPT-PHARMACOMETRICS & SYSTEMS PHARMACOLOGY, 2021. 10(1): p. 67–74.

15. Khurshid, S., et al., ECG-Based Deep Learning and Clinical Risk Factors to Predict Atrial Fibrillation. CIRCULATION, 2022. 145(2): p. 122–133.

16. Ding, H., et al., Evaluating Prognostic Value of Dynamics of Circulating Lactate Dehydrogenase in Colorectal Cancer Using Modeling and Machine Learning. Clinical pharmacology and therapeutics, 2023.

17. Socinski, M.A., et al., Atezolizumab for First-Line Treatment of Metastatic Nonsquamous NSCLC. NEW ENGLAND JOURNAL OF MEDICINE, 2018. 378(24): p. 2288–2301.

18. Kittler, J. and P.C. Young, NEW APPROACH TO FEATURE SELECTION BASED ON KARHUNEN-LOEVE EXPANSION. PATTERN RECOGNITION, 1973. 5(4): p. 335–352.

19. Mercer, J., Functions of positive and negative type, and their connection with the theory of integral equations. PROCEEDINGS OF THE ROYAL SOCIETY OF LONDON SERIES A-CONTAINING PAPERS OF A MATHEMATICAL AND PHYSICAL CHARACTER, 1909. 83(559): p. 69–70.

20. Cox, D.R., REGRESSION MODELS AND LIFE-TABLES. JOURNAL OF THE ROYAL STATISTICAL SOCIETY SERIES B-STATISTICAL METHODOLOGY, 1972. 34(2): p. 187–220.

21. Therneau, T.M. A package for survival analysis in R. R package version 3.1-12. Accessed January 15, 2021; Available from: https://cran.r-project.org/web/packages/survival/vignettes/survival.pdf.

22. Hothorn, T., K. Hornik, and A. Zeileis, Unbiased recursive partitioning: A conditional inference framework. JOURNAL OF COMPUTATIONAL AND GRAPHICAL STATISTICS, 2006. 15(3): p. 651–674.

23. Hothorn, T., et al., Bagging survival tree. STATISTICS IN MEDICINE, 2004. 23(1): p. 77–91.

24. Strobl, C., et al., Bias in random forest variable importance measures: Illustrations, sources and a solution. BMC BIOINFORMATICS, 2007. 8: p. 5.

25. Uno, H., et al., On the C-statistics for evaluating overall adequacy of risk prediction procedures with censored survival data. STATISTICS IN MEDICINE, 2011. 30(10): p. 1105–1117.

26. He, S.S., et al., Dynamic changes in plasma Epstein-Barr virus DNA load during treatment have prognostic value in nasopharyngeal carcinoma: a retrospective study. CANCER MEDICINE, 2018. 7(4): p. 1110–1117.

27. Leung, S.F., et al., Plasma Epstein-Barr viral DNA load at midpoint of radiotherapy course predicts outcome in advanced-stage nasopharyngeal carcinoma. ANNALS OF ONCOLOGY, 2014. 25(6): p. 1204–1208.

28. Dumont, C., et al., PFIM 4.0, an extended R program for design evaluation and optimization in nonlinear mixed-effect models. COMPUTER METHODS AND PROGRAMS IN BIOMEDICINE, 2018. 156: p. 217–229.

29. Davidian, M. and D.M. Giltinan, Nonlinear models for repeated measurement data: An overview and update. JOURNAL OF AGRICULTURAL BIOLOGICAL AND ENVIRONMENTAL STATISTICS, 2003. 8(4): p. 387–419.

30. Gao, C.X., et al., Self-attention-based time-variant neural networks for multi-step time series forecasting. NEURAL COMPUTING & APPLICATIONS, 2022. 34(11): p. 8737–8754.

31. Wang, X., et al., Long Time Series Deep Forecasting with Multiscale Feature Extraction and Seq2seq Attention Mechanism. NEURAL PROCESSING LETTERS, 2022. 54(4): p. 3443–3466.

32. Fan, J., et al., Parallel spatio-temporal attention-based TCN for multivariate time series prediction. NEURAL COMPUTING & APPLICATIONS, 2023. 35(18): p. 13109–13118.

